# Building an International Consortium for Tracking Coronavirus Health Status

**DOI:** 10.1101/2020.04.02.20051284

**Authors:** Eran Segal, Feng Zhang, Xihong Lin, Gary King, Ophir Shalem, Smadar Shilo, William E. Allen, Yonatan H. Grad, Casey S. Greene, Faisal Alquaddoomi, Simon Anders, Ran Balicer, Tal Bauman, Ximena Bonilla, Gisel Booman, Andrew T. Chan, Ori Cohen, Silvano Coletti, Natalie Davidson, Yuval Dor, David A. Drew, Olivier Elemento, Georgina Evans, Phil Ewels, Joshua Gale, Amir Gavrieli, Benjamin Geiger, Iman Hajirasouliha, Roman Jerala, Andre Kahles, Olli Kallioniemi, Ayya Keshet, Gregory Landua, Tomer Meir, Aline Muller, Long H. Nguyen, Matej Oresic, Svetlana Ovchinnikova, Hedi Peterson, Jay Rajagopal, Gunnar Rätsch, Hagai Rossman, Johan Rung, Andrea Sboner, Alexandros Sigaras, Tim Spector, Ron Steinherz, Irene Stevens, Jaak Vilo, Paul Wilmes, CCC (Coronavirus Census Collective)

## Abstract

Information is the most potent protective weapon we have to combat a pandemic, at both the individual and global level. For individuals, information can help us make personal decisions and provide a sense of security. For the global community, information can inform policy decisions and offer critical insights into the epidemic of COVID-19 disease. Fully leveraging the power of information, however, requires large amounts of data and access to it. To achieve this, we are making steps to form an international consortium, Coronavirus Census Collective (CCC, coronaviruscensuscollective.org), that will serve as a hub for integrating information from multiple data sources that can be utilized to understand, monitor, predict, and combat global pandemics. These sources may include self-reported health status through surveys (including mobile apps), results of diagnostic laboratory tests, and other static and real-time geospatial data. This collective effort to track and share information will be invaluable in predicting hotspots of disease outbreak, identifying which factors control the rate of spreading, informing immediate policy decisions, evaluating the effectiveness of measures taken by health organizations on pandemic control, and providing critical insight on the etiology of COVID-19. It will also help individuals stay informed on this rapidly evolving situation and contribute to other global efforts to slow the spread of disease.

In the past few weeks, several initiatives across the globe have surfaced to use daily self-reported symptoms as a means to track disease spread, predict outbreak locations, guide population measures and help in the allocation of healthcare resources. The aim of this paper is to put out a call to standardize these efforts and spark a collaborative effort to maximize the global gain while protecting participant privacy.

## Introduction

The rapid and global spread of COVID-19 led the World Health Organization (WHO) to declare a pandemic on March 11, 2020. One contributing factor to the explosion in cases is the lack of information about who is infected, in large part because not enough people are being tested. This is further compounded in many countries, such as the United States, by health privacy laws. In countries that have tested more aggressively and transparently shared this data, such as South Korea and Singapore, the spread of disease has been greatly slowed ^1^. Although efforts are underway around the world to substantially ramp up testing capacity, technology-driven approaches to collect self-reported information can fill an immediate need and complement official diagnostic results. This type of approach has been used for tracking other diseases, notably flu (see flunearyou.org/covidnearyou.org) ^2^. The voluntary collection of privacy-protected information about individuals’ health status over time will enable researchers to leverage this data to predict, respond to, and learn about the spread of COVID-19. Given the global nature of the disease and our society, we aim to form an international consortium, tentatively named Coronavirus Census Collective, to serve as a hub for amassing this type of data and create a unified platform for global epidemiological data collection and analysis.

### The Mission of the CCC

The CCC is committed to a mission of saving lives through the open sharing of information to the greatest extent possible while simultaneously ensuring privacy. This infrastructure can immediately help in the current COVID-19 pandemic and will also be useful for other diseases that may emerge in the future, or are currently present. Although we are optimistic that capacity for diagnostic testing will rapidly increase, testing will likely never provide global population-wide coverage and there is thus a critical and immediate need for collecting additional data on self-reported symptoms and health status at a population level. Moreover, we plan to integrate the growing official diagnostic testing data along with other real-time informative data to better estimate the symptoms that characterize patients who are diagnosed with COVID-19 and to improve our computational models. In the long term, a broad surveying of individual health status will serve as a rich source of information for understanding disease outbreaks that can guide policy decisions and ensure that the world is better positioned to respond to future pandemics. Nature has presented us with a problem of unprecedented scale that knows no borders. Now is the time to respond with our own global solution: information. Working together, we can act decisively to maximize human health, both now and in the future.

### The Need for Information

Early epidemiological studies of the COVID-19 outbreak in Hubei Province, China, where COVID-19 is thought to have originated from, demonstrated the overwhelming importance of slowing the rate of transmission to reduce the spread of COVID-19. Slowing the rate of transmission, requires information about who is infected and where they are located. China achieved this through testing large numbers of individuals suspected of being infected and moving individuals who tested positive to isolation. In South Korea, which saw the second large spike in COVID-19, government officials have taken an approach of combining large-scale testing with transparent data sharing on where affected individuals are located. Although the success of this approach is clear, it has not been universally implemented. For example, Italy has not adopted this approach, and their rates of infection have steeply risen in recent days, with lack of universal testing potentially playing a role in this unfortunate development. While China’s and South Korea’s approach has been successful, it has two major limitations that prevent its application in the vast majority of other countries: diagnostic testing capacity is limited and personal and health privacy laws are more restrictive. For example, in the United States, an emerging world epicenter, large-scale testing is still not available in the vast majority of US states. For several weeks after community spreads have been documented in multiple US states, the tests were still limited to people with severe symptoms and with international travel history to places with early outbreak (e.g. China, South Korea, Japan, Iran). With limited capacity in testing, the numbers reported as “confirmed cases” in our communities (e.g. https://coronavirus.jhu.edu/) do not reflect the true numbers or the actual rate of COVID-19 spread. The fact that many carrier individuals only show mild symptoms and not seeking tests also contributes to our lack of understanding of the impact.

Technology-driven approaches to collect voluntarily self-reported data on health status can overcome these limitations. These can be further integrated with other relevant real-time data resources such as meteorological data, population density at a given time and place, as well as other dynamic data sources. Together these can provide crucial information that can be immediately leveraged for early identification of disease clusters, with the goal of slowing the spread of disease.

We started several such efforts in the U.S. and one in Israel. In the U.S., we have developed an app, HowWeFeel (http://www.howwefeel.org), that administers a 30-second survey of the person’s well-being to collect epidemiological data (Figure 1). Independently, CovidNearYou was started (https://covidnearyou.org/) as well as covid19-EIPM, a web-application with open-access visualization tools collecting the same information in a fully anonymized way (https://covid19.eipm-research.org) (Figures 3 and 4). In Israel, we developed Predict-Corona (https://coronaisrael.org/), a daily one-minute online survey in which respondents are asked to measure body temperature and report whether they experience any of the symptoms that were found to be common in patients with COVID-19 infection according to the existing literature. Within approximately 10 days of its launch, there have been over 300,000 responses, and we are already seeing the potential of this approach for detecting future outbreak regions (Figure 2) 3.

**Figure 1:**
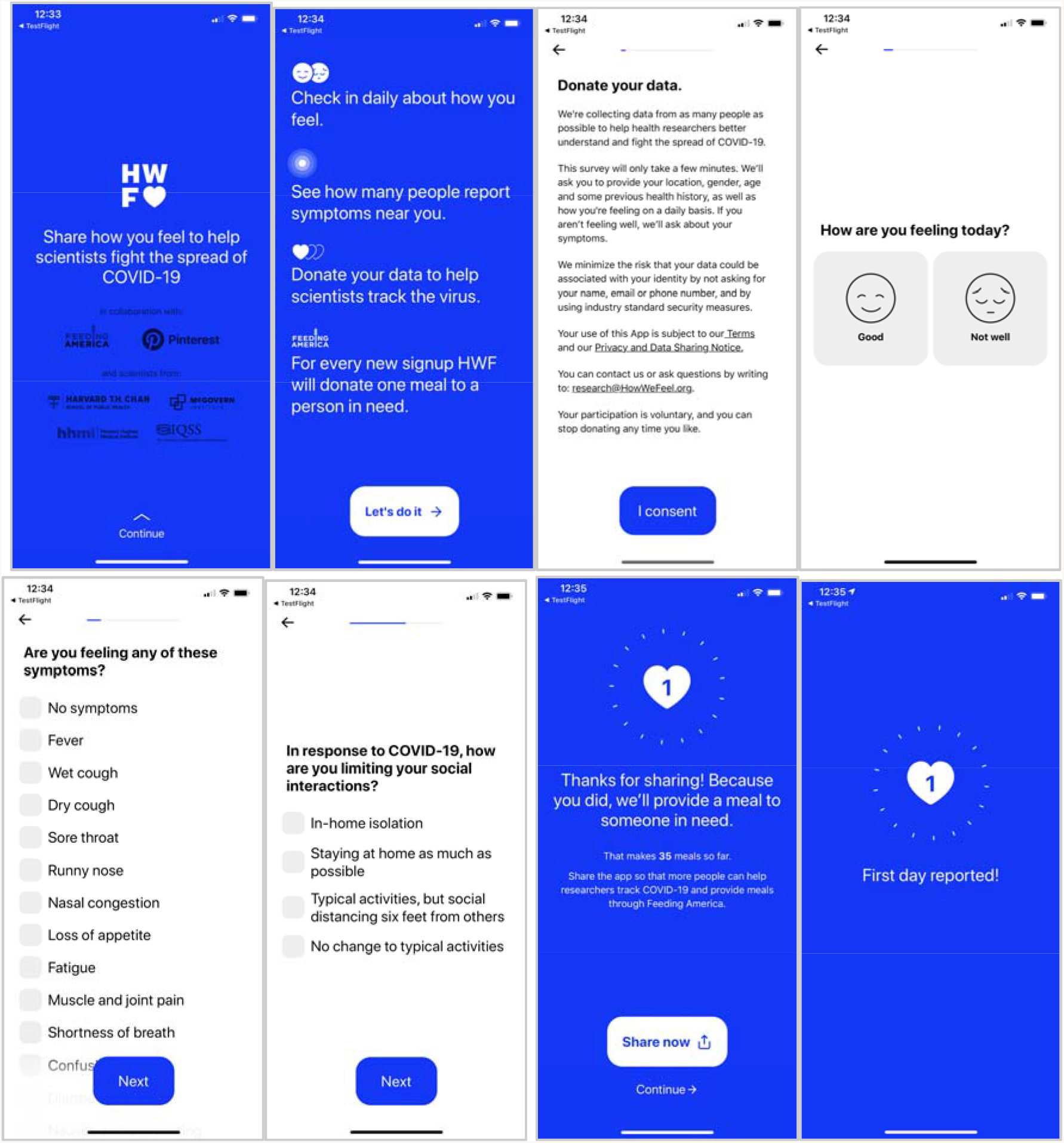
HowWeFeel: a 30-second survey of the person’s well-being to collect epidemiological data.

**Figure 2:**
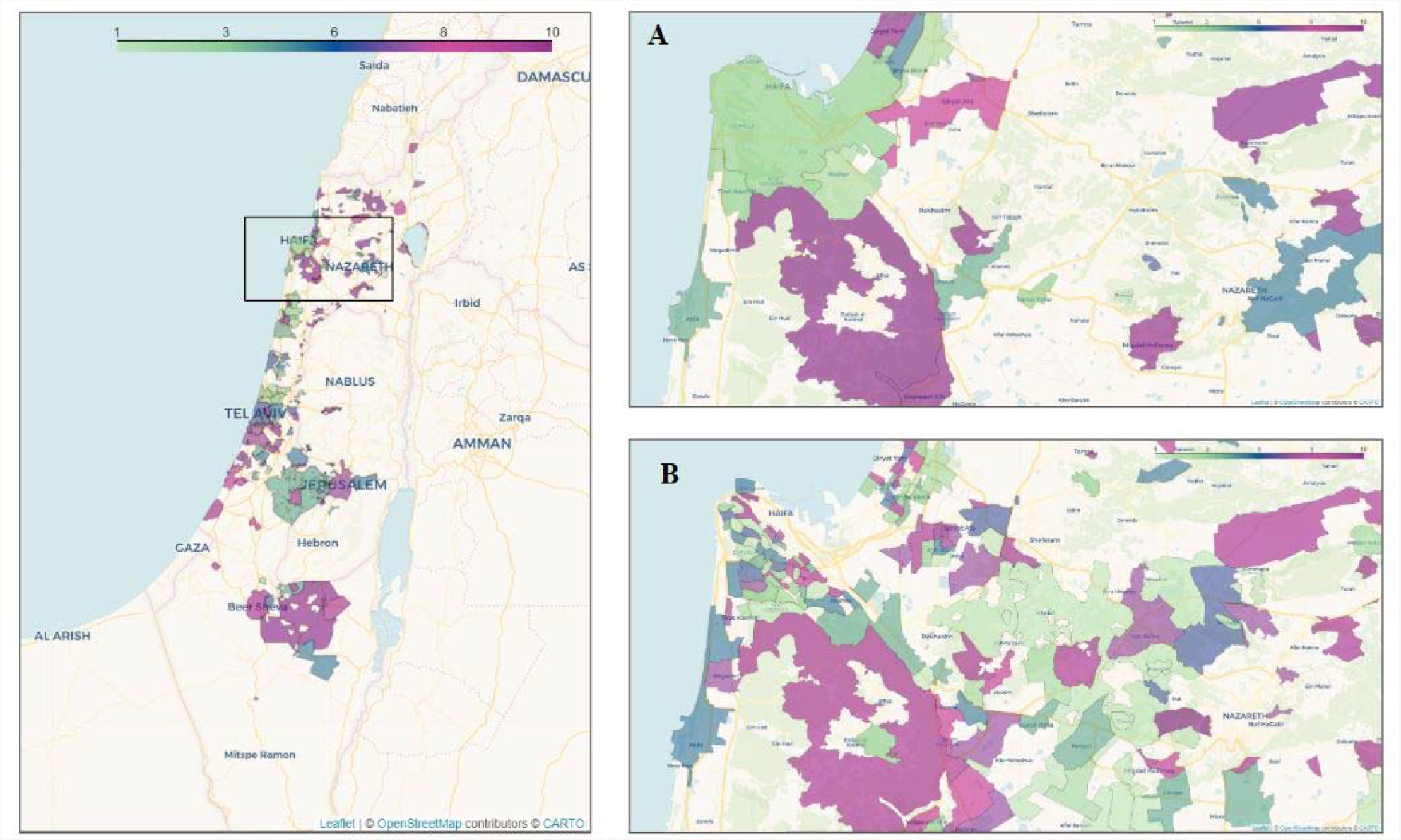
Average COVID-19 associated symptoms region map in Israel. City municipal regions with at least 50 responders and neighborhoods with at least 15 responders are shown. Each region is colored by a category defined by the average symptoms ratio, calculated by averaging the reported symptoms rate by responders in that city or neighborhood. Green - low symptoms rate, Purple - high symptoms rate. **A:** Area of Haifa with city regions. **B:** Area of Haifa with neighborhood regions. For updated maps see https://coronaisrael.weizmann.ac.il/

**Figure 3:**
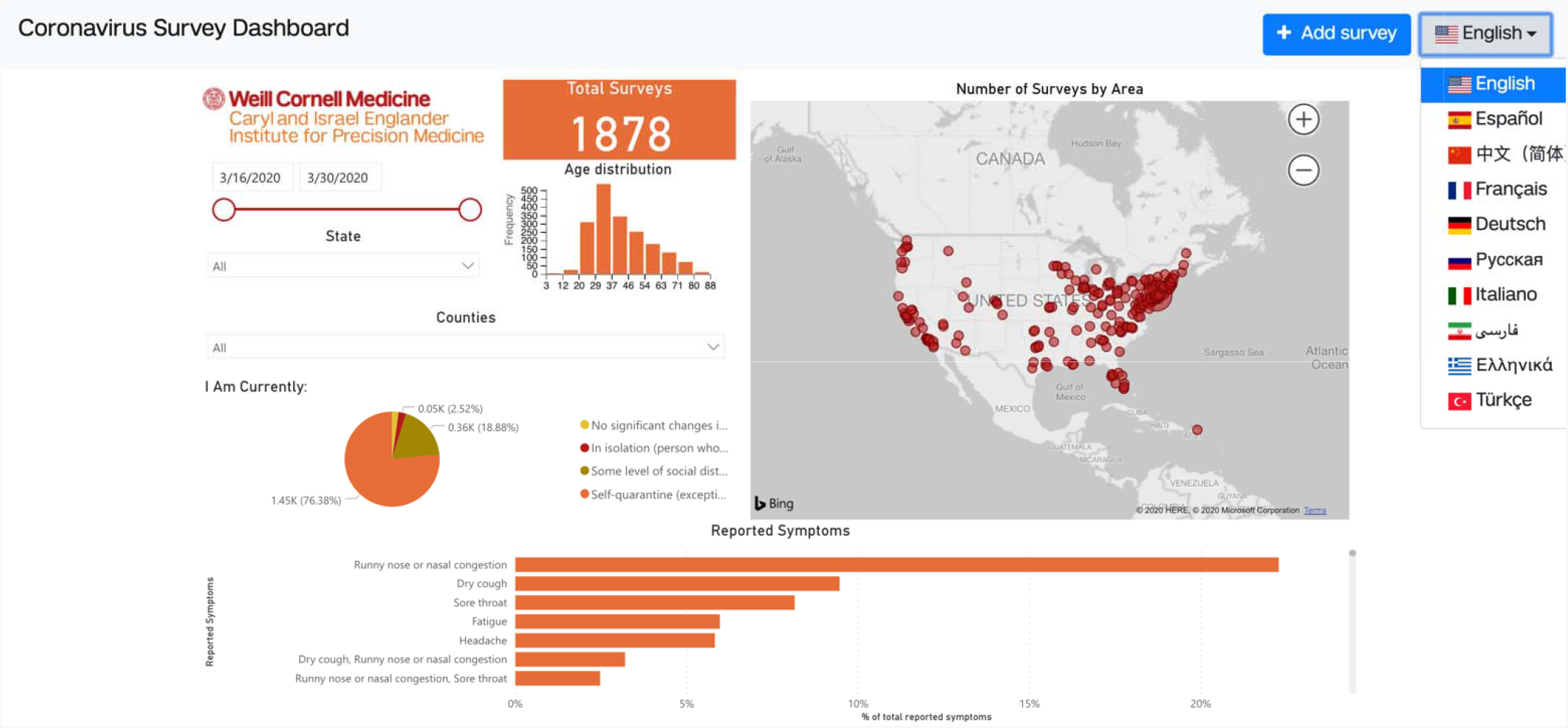
The screenshot of EIPM Coronavirus Survey Dashboard taken on 30/3/2020 is shown in this figure. The dashboard shows collected results at the time a user accesses our website and several visualization tools enable onlin analysis. Statistics on reported symptoms at the state and county level can be obtained from the dashboard. Users can participate using the “Add survey” button. Currently, this allows choosing between 10 different languages to reflect th diversity of our users.

**Figure 4:**
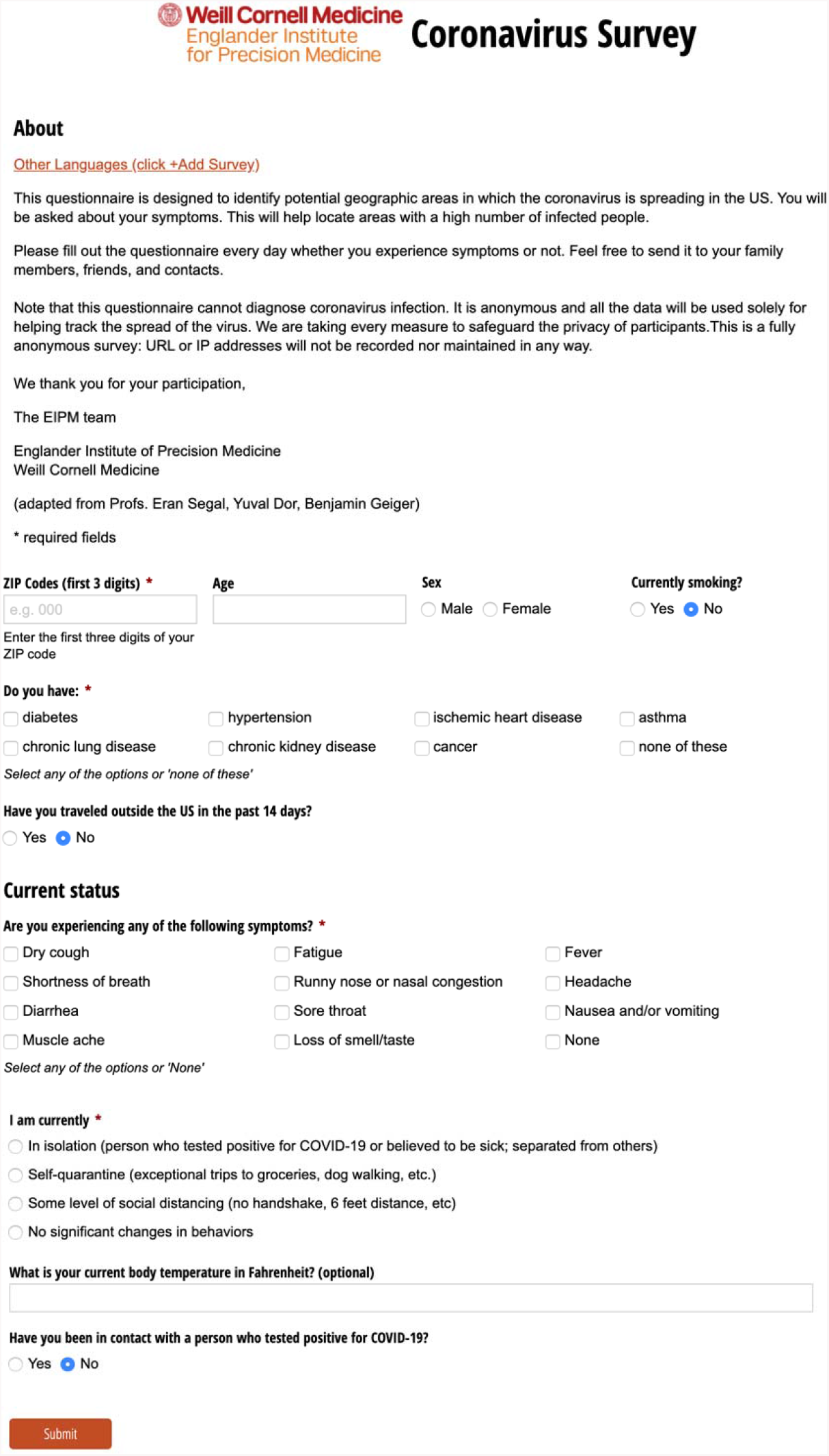
The English version of the EIPM daily questionnaire. Required questions are marked with (*). In particular, the user is required to enter the first three digits of their US zip code. This allows tracking of symptoms in different geographical locations in the entire country. In order to retain HIPAA Privacy Rule compliance, zip codes with less than 20,000 individuals were excluded.

This work parallels efforts in other countries to develop similar surveys, such as the Covid Symptom Tracker in the UK (https://covid.joinzoe.com/), the COVID-19: CH Survey Project in Switzerland (https://covid19survey.ethz.ch/) and Covid-19 self-reporting in Slovenia (https://covid-19-stats.si/). In the U.S. and UK., we have established the COronovirus Pandemic (COPE) Consortium (https://www.monganinstitute.org/cope-consortium) to use the Covid Symptom Tracker (https://covid.joinzoe.com) to join the efforts of international prospective and observational cohorts (e.g. population based, clinical, etc.) and clinical trials/studies.

To increase the statistical power of these data, we call for the formation of an international consortium, the Coronavirus Census Collective (CCC), to integrate these data streams and provide a single, central data bank that researchers from around the world can query securely. In addition to ensuring comparability in the data, the CCC will serve as a resource for entities in other countries that are developing such surveys with the goal of facilitating its rapid deployment around the world. As many countries are now struggling to find the best strategy to handle the pandemic, we believe that a global collaborative effort to obtain data that can be used to predict outbursts of COVID-19 infection is urgently needed.

In carrying out this global endeavor, we acknowledge the importance of catering both to underdeveloped countries and to populations that are underrepresented or of lower socio-economic status. For example, in Israel we distributed our survey in several languages, engaged leaders in local religious communities, and promoted the survey through both Hebrew and Arabic-speaking media channels to increase compliance across all sectors of the Israeli population. Similarly, HowWeFeel has been translated into 20 languages and covid19-EIPM currently supports 10 different languages.

### A Framework for Collecting and Sharing Data

We will collect survey responses containing symptomatic information relevant to COVID-19 along with geospatial location, time, and demographic information on age and pre-existing and comorbid medical conditions. The data will be collected, de-identified, and further made differentially private before analysis to enable researchers to share and analyze the data while maintaining individual level privacy Because the privacy of our participants is essential to our mission, we will remain at the cutting edge of privacy protective technologies, develop novel methods where needed, and implement improvements over time wherever possible. Our technologies and the code for our app will be open source, so others adapt for their particular situations, find bugs, or help us improve it. We envision that all members will use surveys with a common set of “core” questions, but additional region-specific questions may be added within the extent of the local regulations, researcher interest, and community need. This set of core questions may grow over time as more is learned about COVID-19.

A federated common data model will be built to facilitate data sharing while ensuring data security and privacy across different countries. To achieve that goal, we will define guidelines to ensure that the underlying data model collected from all contributors can easily be amalgamated and harmonized, and the consortium data sharing policy will be developed. We will also apply different methods such as differential privacy ^4^ which will enable researchers to share and analyze the data while preserving the confidentiality of the participants. Given differences in privacy regulations from region to region, individual level data from the surveys from some countries may not be accessible, but the results of differentially private statistical analyses will be available to all.

These data will allow researchers to perform several immediately useful analyses. For example, statistics can be computed of how many individuals exhibit symptoms, or self-report official COVID-19 test results within particular geographic regions. Statistics can be computed both in aggregate and conditional on comorbidities and demographic information such as age when available. This will enable monitoring of the health status of the respondent population, either in aggregate or stratified by region or demography, at any given time. Regression analysis will be performed to investigate the epidemiological factors associated with symptoms and testing results. Spatial analysis will be performed and geostatistical maps on the prevalence of symptoms and positive test probability will be constructed. In addition, clustering models can be used to identify which areas likely have COVID-19 outbreaks already ongoing based on co-occurrence of many individuals reporting similar patterns of symptoms at the same time. This will enable estimates of the population of individuals who have COVID-19 symptoms in spite of inadequate viral testing. With participants who have tested positive and those who tested negative, we will be able to use methods of “quantification” (as distinct from “classification”) to produce accurate estimates of population prevalence of COVID-19 even when individuals cannot be reliably classified from their symptoms alone ^5^. Another possible application is evaluating the effectiveness of the various social distancing measures taken, and their contribution to reduction of the number of symptomatic people. Finally, lagged prediction models can be used to identify increasing trends in the incidence of self-reported COVID-19 related symptoms within specific geographic locales. These predictions will be potentially useful for identifying areas where additional testing or medical resources should be allocated. To maximize the impact of the data collected, we will provide an Application Programming Interface (API) to allow accredited researchers to access the data to perform additional statistical analyses.

The CCC aims to further leverage the impact of voluntary self-reported health information by actively seeking collaboration from others around the world that are similarly committed to maximizing the potential benefits of this approach while maintaining individual’s privacy. We envision that the CCC will be coordinated by a Board of Directors, who will vet new members, maintain a secured centralized data repository, or federated multi-site data repositories, and develop mechanisms to enable researchers from around the world to query the data. Individual members will be responsible for ensuring adherence to local regulations.

## Data Availability

The CCC is a non-profit consortium open to anyone who shares the same vision of making data available to help the public good and fight COVID-19. There are no membership fees. Please contact us at if you are interested in joining.

